# Interdependence between confirmed and discarded cases of dengue, chikungunya and Zika viruses in Brazil: A multivariate time-series analysis

**DOI:** 10.1101/19002246

**Authors:** Juliane F Oliveira, Moreno S. Rodrigues, Lacita M. Skalinski, Aline ES Santos, Larissa C. Costa, Luciana L. Cardim, Enny S. Paixão, Maria da Conceição N. Costa, Wanderson K. Oliveira, Maurício L. Barreto, Maria Glória Teixeira, Roberto F. S. Andrade

## Abstract

The co-circulation of different arboviruses in the same time and space poses a significant threat to public health given their rapid geographic dispersion and serious health, social, and economic impact. Therefore, it is crucial to have high quality of case registration to estimate the real impact of each arboviruses in the population. In this work, a Vector Autoregressive (VAR) model was developed to investigate the interrelationships between discarded and confirmed cases of dengue, chikungunya, and Zika in Brazil. We used data from the Brazilian National Notifiable Diseases Information System (SINAN) from 2010 to 2017. There were two waves in the series of dengue notification in this period, one occurring in 2013 and the second in 2015. The series of reported cases of both Zika and chikungunya reached their peak in late 2015 and early 2016. The VAR model shows that the Zika series have a significant impact on the dengue series and vice versa, suggesting that several discarded and confirmed cases of dengue could actually have been cases of Zika. The model also suggests that the series of confirmed chikungunya cases is almost independent of the cases of dengue and Zika. In conclusion, co-circulation of arboviruses with similar symptoms could lead to misdiagnosed diseases in the surveillance system. We argue that the use of mathematical and statistical models routinely in association with traditional symptom-surveillance could help to decrease such errors and to provide early indication of possible future outbreaks. These findings address the challenges regarding notification biases and shed new light on how to handle reported cases based only in clinical-epidemiological criteria when multiples arboviruses co-circulate in the same population.

**Author summary:** Arthropod-borne viruses (arboviruses) transmission is a growing health problem worldwide. The real epidemiological impact of the co-circulation of different arboviruses in the same urban spaces is a recent phenomenon and there are many issues to explore. One of this issue is the misclassification due to the scarce availability of confirmatory laboratory tests. This establishes a challenge to identify, distinguish and estimate the number of infections when different arboviruses co-circulate. We propose the use of multivariate time series analysis to understand how the weekly notification of suspected cases of dengue, chikungunya and Zika, in Brazil, affected each other. Our results suggest that the series of Zika significantly impact on the series of dengue and vice versa, indicating that several discarded and confirmed cases of dengue might actually have been Zika cases. The results also suggest that the series of confirmed cases of chikungunya are almost independent of those of dengue and Zika. Our findings shed light on yet hidden aspects on the co-circulation of these three viruses based on reported cases. We believe the present work provides a new perspective on the longitudinal analysis of arboviruses transmission and call attention to the challenge in dealing with biases in the notification of multiple arboviruses that circulate in the same urban environment.

## Introduction

In recent times, the re-emergence and the rapid spread of arboviruses in urban areas have become a serious problem that has concerned health authorities as well as the general population in many countries. The magnitude of the epidemics, the occurrence of severe cases with neurological manifestations and lethal outcomes, and severity of congenital malformations associated with infections occurred during pregnancy are the main threats of this new epidemiological situation [1, 2].

In Brazil, the co-circulation of the four serotypes of dengue virus (DENV), together with the emergence and dissemination of chikungunya virus (CHIKV) and Zika virus (ZIKV), transmitted by *Aedes* mosquitoes (mainly *Aedes aegypti*), has a relevant negative impact on the health of the population and lead to an increase in the demand on health and other support services. From their introduction, in 1986, to until arrival and subsequent spread of CHIKV and ZIKV, DENV was the most important vector-borne disease circulating in cities of Brazil, [3, 4]. In September of 2014, CHIKV was detected in cities of the states of Amapá and Bahia. Although this disease causes arthralgia with pain at a higher level than dengue, the other symptoms are similar, which increased the likelihood of misdiagnosis [5]. In October 2014, an outbreak of an undetermined exanthematous illness was registered in Rio Grande do Norte, in the northeast of Brazil, when in April 2015, ZIKV was identified as the aetiologic agent [6]. Patients infected with ZIKV typically presented low (or no) fever and skin rash within the first 24 hours of the disease onset, while DENV and CHIKV cause high fever immediately. Also, CHIKV causes more intense arthralgia than DENV. However, the other symptoms are similar, which confound and complicate their differentiation and easily lead to misdiagnosis, [2, 7, 8].

The similarity of symptomatology has made the clinical diagnosis of arboviruses difficult, especially in the course of epidemics with viral co-circulation, in which laboratory tests are still unavailable for most patients. The misclassification and incorrect diagnosis affect the risk estimates of these diseases since epidemiological surveillance depends on the quality of the data to provide morbidity and mortality information close to the reality lived by the population and, consequently, the development of effective prevention strategies, [2, 7, 9]. Therefore, this study aims to explore and understand how dengue notified cases were impacted by the introduction and spread of chikungunya and Zika virus in Brazil.

## Materials and methods

We used a multivariate time series analysis in order to understand how the classification of notified cases of dengue, Chikungunya and Zika interact with each other in Brazil from 2015 to 2017.

### Data source

We use data from the Brazilian National Notifiable Diseases Information System (SINAN). We collected weekly reported data of suspected cases of dengue (from 2010 to 2017), chikungunya (from 2014 up to 2017), and Zika (from 2015 up to 2017) viruses. We only considered cases that presented non-null information about the temporal variable, i.e., week of notification or week of first symptoms. We further separated the cases into confirmed and discarded, following the final classification information as used by the Brazilian Ministry of Health. Confirmed cases are all suspected cases of disease, excluding those discarded or inconclusive. This classification can be based on clinical/epidemiological criteria, namely presence of clinical symptoms in the same area and time as other probable cases, or on clinical/laboratory criteria, namely the presence of clinical symptoms and a positive IgM ELISA result, viral RNA detection via PCR, NS1 viral antigen detection, or positive viral culture [3]. Discarded cases are defined as any suspected case that satisfies at least one of the following criteria: negative laboratory diagnosis (IgM serology); a laboratory confirmation of another disease; clinical and epidemiological compatibility with other diseases. Inconclusive cases of dengue and Chikungunya were excluded from the analyses. However inconclusive cases of Zika represented about 30% (110,656/361,396 registered cases) of all notified cases from 2015 to 2017, accounting for about 56% (33,863/60,972 registered cases) of the Zika notifications in 2015. Therefore, for the purpose of this study, we considered inconclusive Zika cases as probable Zika cases.

To perform the study of time series analyses we collected the confirmed and discarded reported cases of each arbovirus per epidemiological week in Brazil, from 2015 to 2017.

### Multivariate time-series analysis

We construct a Vector Autoregressive (VAR) model to uncover possible correlation and causality effects between the discarded and confirmed cases of the three arboviruses.

Formally, a time-series is defined as *random sequence*, i.e., a collection of random variables {*Y*_*t*_}, where the time-index assumes integer values only. A univariate time-series {*Y*_*t*_} is said to be an *autoregressive process* if each *Y*_*t*_ is defined in terms of its predecessors *Y*_*p*_, for *p* < *t*, by the equation:

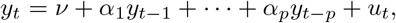

where *ν* is a fixed constant of intercept terms allowing to the possibility of a non-zero mean, and {*U*_*t*_} is a *white noise*, i.e., a sequence of mutually independent random variables, each with mean zero and finite variance *σ*^2^.

If a *m*-dimensional multivariate time-series is considered, a vector autoregressive process (VAR) is defined as a generalization of definition of autoregressive process given by:

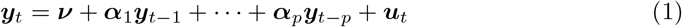

where now the bold letters represent a vector notation. Thus, ***Y***_*t*_ = (*Y*_1*t*_, …, *Y*_*mt*_), ***ν*** = (*ν*_1_, …, *ν*_*m*_) are *m*-vector of constants, ***α***_1_, …, ***α***_*p*_ forms a (*m* × *p*)-matrix of coefficients, and ***U***_*t*_ is a multivariate *m*-vector white noise.

We say that (1) is a VAR process of order *p*, VAR(*p*), if ***α***_1*p*_, …, ***α***_*mp*_ ≠ 0 and ***α***_1*i*_, …, ***α***_*mi*_ = 0 for *p* < *i*, where *p* is the smallest possible order.

In this work, we consider a 6-dimensional multivariate time series, with each *Y*_*t*_ representing a vector (*Z*_1*t*_, *Z*_2*t*_, *C*_1*t*_, *C*_2*t*_, *D*_1*t*_, *D*_2*t*_), where *Z, D*, and *C* denotes Zika, dengue and Chikungunya respectively, the indices 1 and 2 stand for confirmed and discarded cases, and the time *t* ranges from the first week of 2015 until the last week of 2017.

The steps to construct and analyse a VAR(*p*) model consist of: (i) setting the *p* (lag) order, which is is automatically selected by the minimum of Akaike information criterion (AIC); (ii) estimation of the VAR coefficients by a multivariate Least Squares Estimation; (iii) test for normality of residuals, using a probability plot to assess how the residual error depart from normality visually, and an analysis of the partial (cross-)correlation function (PCF) between them; (iv) perform Granger test for causality for the 6-dimensional estimated multivariate series.

Using the Granger causality F-test in a pair-wise way, we can check whether the null hypothesis, stating that one series {*X*_*t*_} does not affect the other one {*Y*_*t*_}, can be rejected or not. If the hypothesis is rejected, then the time-series {*X*_*t*_} Granger-causes a time-series {*Y*_*t*_}. Thus, the past values of {*X*_*t*_} can be used for the prediction of future values of {*Y*_*t*_}. In other words, the values used for describing the autoregressive equation for {*Y*_*t*_} have significant non-zero contribution of past values of {*X*_*t*_}.

In the current context, when a series of discarded cases of disease 1 affects the confirmed cases of disease 2, possibly there is evidence that individuals truly infected by disease 2 were wrong notified as disease 1. However, when confirmed cases of disease 1 affect the discarded cases of disease 2, this can be interpreted as an increase (or decrease) in the notification of disease 2, but not necessarily this notification could be claim as a confirmed case of disease 1.

In order to carry out the analysis, we first rand the vector series {*Y*_*t*_} to a stationary form, in such a way that its mean value and the covariance among its observations do not change with time. Detailed information about the theoretical background for time series analysis can be found in [10, 11].

We performed our statistical analysis using a Python software [12].

### Ethics Statements

We obtained ethics approval from the Federal University of Bahia research ethics committee, Salvador, Brazil (CAAE: 70745617.2.0000.5030). All data analyzed were anonymized

## Results

From January 2010 to December 2017, it was registered in SINAN 9,936,488 million cases of dengue, from which 56% (5,570,157/9,936,488) were confirmed and 32% (3,138,750/9,936,488) were discarded. From January 2014 to December 2017 it was registered 501,202 thousands of cases of chikungunya, resulting in 63% (317,158/501,202) and 27% (128,100/501,202) of confirmed and discarded cases, respectively. The classified confirmed and discarded cases of Zika, from January 2015 to 2017, represented 76% (286,198/375,251) and 24% (89,053/375,251) of registered cases, respectively.

During the period covered by this survey, the confirmed cases of dengue had its peak in early 2015, although the worst epidemy of the disease occurred in Brazil was registered in 2013. Confirmed Zika and chikungunya cases reach their peak in late 2015 and middle of 2016 respectively. For a visualization of these traits, see S1 Fig. the Fig 1 shows the curves of confirmed and discarded cases from 2015 to 2017 of the three diseases.

**Fig 1.**
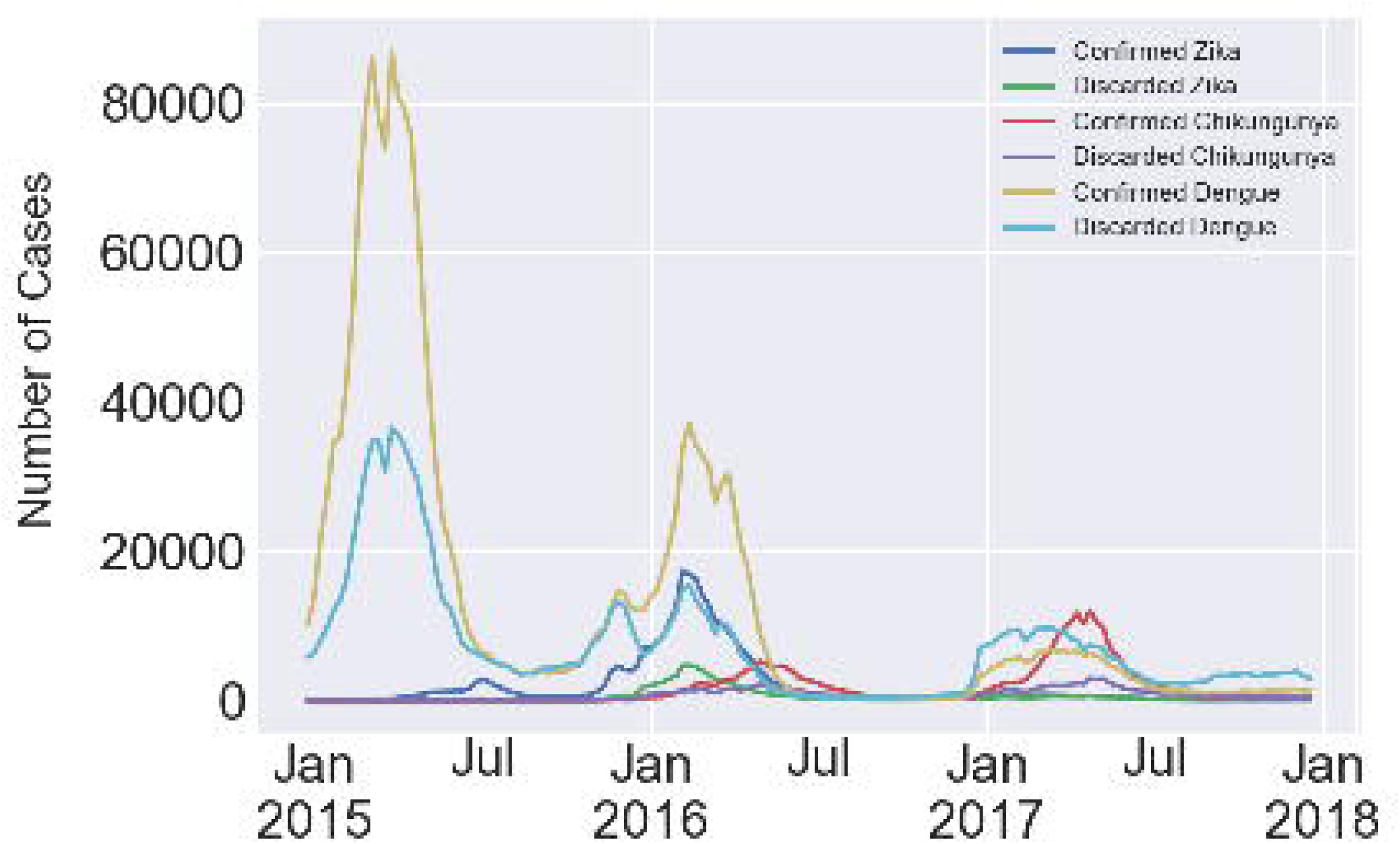
Illustration of time series plots of confirmed and discarded cases of dengue, chikungunya and Zika by epidemiological week. Brazil, January 2015 to December 2017.

Fig 2 depicts a linear dependence between the series of confirmed and discarded cases per epidemiological week, during the whole corresponding periods, for dengue, chikungunya, and Zika in the country. The slope *a*_*D*_ of the linear relation for dengue is 2.4, while yearly based evaluations lead to a mean value ⟨*a*_*D*_⟩_*t*_ = 2.3 (*SD* = 1). It means that, on average, for every ten confirmed cases of dengue, about 4 cases are discarded. For chikungunya and Zika, the slopes are 3.2 and 3.6 respectively.

**Fig 2.**
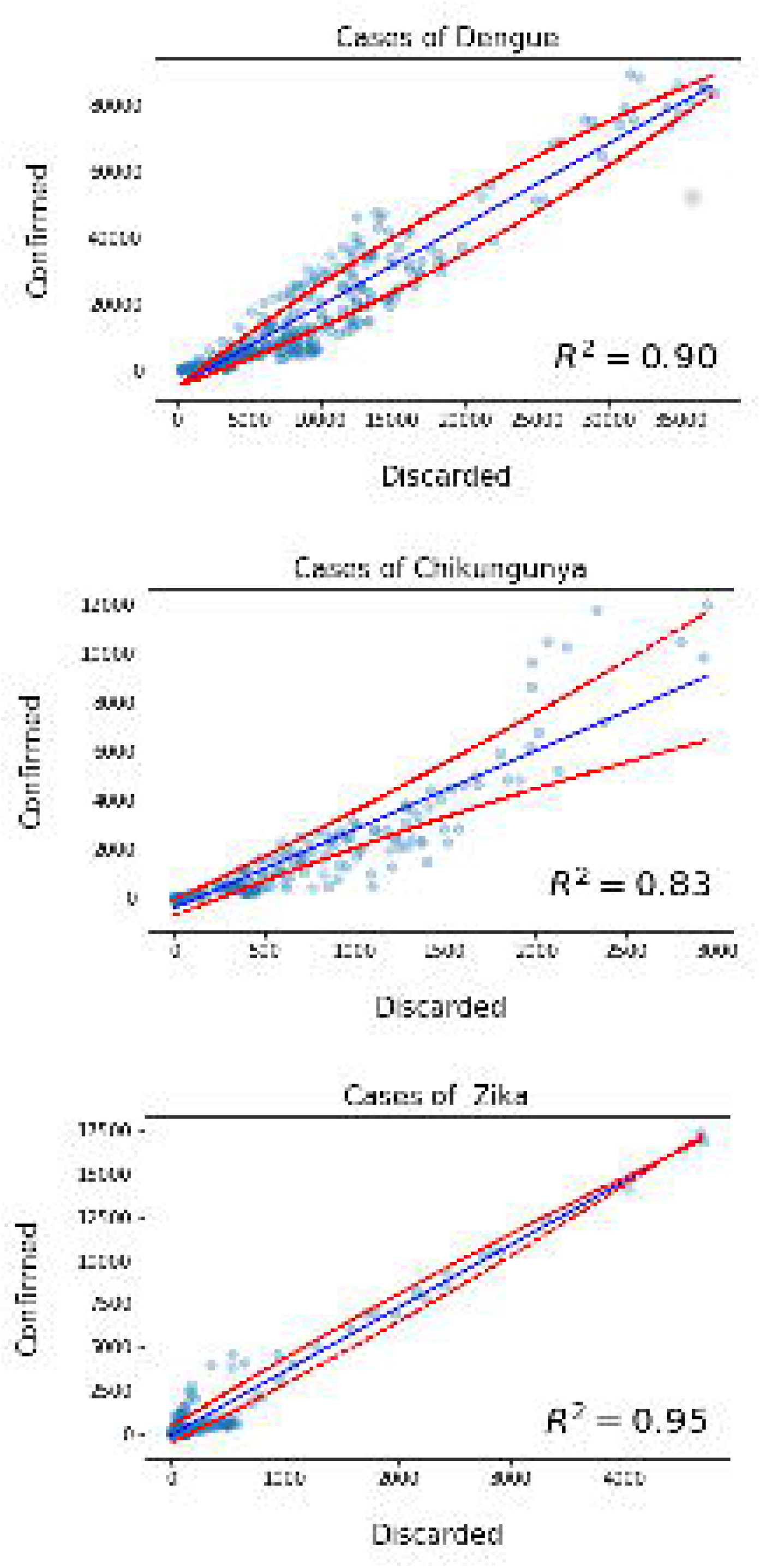
Scatter plot of confirmed against discarded reported cases in Brazil. dengue (from 2010 up to 2017); chikungunya (from 2014 up to 2017); Zika (from 2015 up to 2017). The red dashed lines represent the error interval.

For the multivariate time series of the study, the AIC result for the lag *p* was 13. Table 1 shows the correlation matrix of the stationary series of confirmed and discarded cases of dengue, chikungunya and Zika. We interpret the values of positive/negative correlation according to the interval: ±.00 to ±.10 very low; ±.10 to ±.40 as weak; ±.40 to ±.60 as moderate; ±.60 to ±.80 as strong; ±.80 to ±.99 very strong; ±1.0 as perfect. Summary of regression results are presented in S1 Appendix. The autocorrelation, cross-correlations and probability plots of residuals are given from S2 Fig to S5 Fig.

**Table 1.** Correlation matrix of the stationary series of confirmed and discarded cases of dengue, chikungunya and Zika. Brazil, January 2015 to December 2017.

|  | *Conf.Zika* | *Disc.Zika* | *Conf.chik.* | *Disc.chik.* | *Conf.dengue* | *Disc.dengue* |
| --- | --- | --- | --- | --- | --- | --- |
| *Conf.Zika* |  | .93 | .04 | .17 | .40 | .37 |
| *Disc.Zika* | .93 |  | .10 | .27 | .42 | .35 |
| *Conf.chik.* | .04 | .10 |  | .66 | .05 | .12 |
| *Disc.chik.* | .17 | .27 | .66 |  | .15 | .24 |
| *Conf.dengue* | .40 | .42 | .05 | .15 |  | .83 |
| *Disc.dengue* | .37 | .35 | .12 | .24 | .83 |  |

The results of the Granger tests to explore associations between series are presented in Table 2. They show that, at a significant level, the series of confirmed cases of Zika affects the series of discarded cases of dengue (Test statistic = 2.807, p-value = 0.001), discarded cases of chikungunya (Test statistic = 2.158, p-value = 0.011) and confirmed cases of dengue (Test statistic = 3.222, p-value < 0.001). In the other way around, there is a significant association between discarded cases of dengue and both confirmed cases of Zika (Test statistic = 3.348, p-value < 0.001) and confirmed cases of chikungunya (Test statistic = 3.444, p-value < 0.001). The series of confirmed Zika and discarded dengue presented a positive weak linear correlation (0.37), which was stronger than the other correlations for the series described above.

**Table 2.**
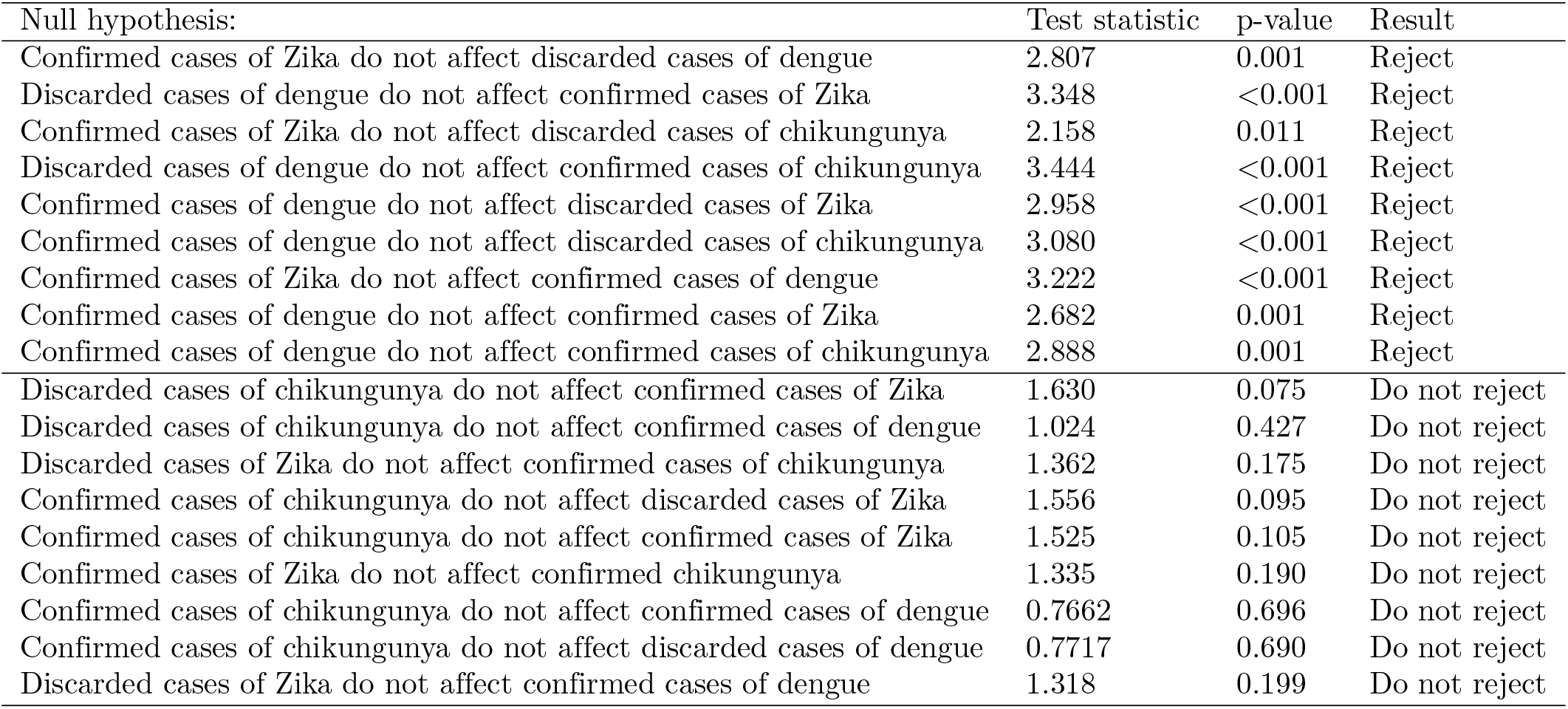
Pairwise Granger causality tests. Exploratory search of associations between series of confirmed and discarded cases of dengue, chikungunya and Zika. Brazil, January 2015 to December 2017.

There is a significant association between confirmed cases of dengue and discarded (Test statistic = 2.958, p-value < 0.001) and confirmed cases of Zika (Test statistic = 2.682, p-value = 0.001). These two series have a positive moderate linear correlation (0.42 and 0.40, respectively). We also found that confirmed cases of dengue have a significant association with discarded (Test statistic = 3.080, p-value < 0.001) and confirmed cases of chikungunya (Test statistic = 2.888, p-value = 0.001). However, by assessing the correlation matrix given in Table 2, the values present a very low correlation (0.15 and 0.05, respectively).

## Discussion

The moderate positive correlation found in the analyses shows that the notification series of dengue was significantly impacted by Zika, and vice-versa. The Granger score only fails to evidence out a causal link between discarded cases of Zika and confirmed cases of dengue. Reasonable interpretation is that an increase of individuals notified as Zika contributes to an increase of wrong notification of dengue cases within two scenario: people with Zika were wrongly classified as dengue (and vice-versa), or perhaps both viruses infected the same individuals. The results also indicate that the series of confirmed Zika cases increased the series of discarded dengue cases and that, among those discarded, there were indeed confirmed Zika cases. The series of confirmed cases of dengue affects significantly and positively (increasing) the series of discarded cases of Zika, which can be explained by the awareness of the consequences accounted by Zika at the end of 2015.

The notifications of Zika and dengue had overall weaker effects on the notification of suspected chikungunya cases, as indicated by the smaller correlation values between the corresponding series. The association of these values with Granger scores indicates possible a causal relationship in 3 of the 12 possible combinations. Considering Zika and chikungunya, it is possible to conclude that the increase of the reported cases of Zika contributed to a meagre increase of suspected chikungunya cases. As to the mutual influence of dengue and chikungunya notifications, Granger causality scores show that possibly a tiny amount of discarded cases of dengue were confirmed cases of chikungunya. They also show that an increase in the series of confirmed dengue cases possibly contributed in a very small increase in the number of discarded cases of chikungunya. In opposition to these three specific combinations, the remaining results show either the failure to reject the null hypothesis, as in the result for discarded chikungunya and confirmed Zika cases, or a very small correlation value, as for both confirmed dengue and chikungunya cases. Thus, the remaining analyses between confirmed and discarded Zika and confirmed chikungunya cases suggest that they did not affect each other and probably their notifications happened as independent events in Brazil. They also support the conclusion that the notifications of dengue and Zika happened independently of the notification of chikungunya.

This study highlighted that in Brazil, from 2015 to 2017, the series of confirmed and discarded cases of dengue, chikungunya and Zika presented, in most of the cases, linear dependence. This reflects the epidemiological context presented by this country from the second semester of 2014 on, when the simultaneous circulation of DENV, ZIKV and CHIKV in densely populated urban spaces greatly hampered the correct record of each case of these diseases [9, 13]. Although CHIKV and ZIKV emerged almost simultaneously in cities in the same region of the country, the latter was only identified at the end of April 2015 [6]. Thus, there was a delay in alerting the health services network about the existence of this new clinical entity. In spite of the long experience in dengue of the professionals of the network of health services of this country, the circumstances presented above did not allow the adequate clinical and epidemiological diagnosis of the cases of each of these three diseases, resulting, often than not, in incorrect records [2].

In a scenario where only the notification of the diseases are available and laboratory tests are scarce, we see that the notification of dengue and Zika are shown to be independent of the notification of chikungunya. These findings are plausible, since dengue and Zika present more similar clinical manifestations to each other as compared to chikungunya [7]. The expressive joint manifestations produced by CHIKV infections allow a more accurate clinical diagnosis, even when specific laboratory tests are not available. Therefore, these results would not support the use of discarded cases of chikungunya as complementary cases of Zika infection, as presented by [13]. However, as the total number of chikungunya discarded cases was small (3.8 %) in comparison to the universe of cases of the three diseases, that fact did not affect the temporal trend presented for this and our study. Nevertheless, it is worth noticing that our results should be nearest of the real, and thus contribute to construct more accurate prediction models of future Zika epidemics, using only possible cases of dengue.

Another point that calls attention is that the lack of association between the series of confirmed cases of dengue and confirmed chikungunya, and confirmed cases of Zika and confirmed chikungunya might be due to spatial factors that are not considered in this work, or to a hypothetical situation where one virus inhibits the proliferation of the other.

Our studies based on a rather simplified linear model can be complemented by future works, where the analysis proceeds either through non-linear methods or through a more comprehensive and adequate model. A detailed study, probably based on the symptoms presented by patients, may also contribute to having better estimations for the quantity of cases that can be assigned for each disease. All suppositions made here are based only on a temporal analysis of the time series of notifications. Therefore, including a spatial analysis would clarify more issues regarding the surveillance of co-circulation of arboviruses and how zones with higher incidences handled with the notifications of the cases.

Overall difficulties regarding inadequate diagnostics and limited availability of laboratory tests for the recent arboviruses outbreak have been pointed out for other authors. However, it is of crucial importance to be able to understand how the arboviruses interact, as this allows to provide a better estimation of the risk and propagation of the disease. At this stage of understanding, we believe that our results raise a discussion of miss-reporting cases and suggest directions for the analysis to assist such a difficulty.

In summary, we demonstrate two important interrelated aspects: the first one refers to how the discarded cases, which resulted from reported cases of one arbovirus, can be considered as part of complementary notifications of another; the second concerns how the series of confirmed cases of one disease may affect the series of confirmed cases of another. Thus, these findings address the challenges regarding notification biases and shed new light on how to handle reported cases based only in criteria clinical-epidemiologic when these three arboviruses co-circulate in the same population.

## Data Availability

All data will e available in the manuscript after acceptance in journal.

## Supporting information

**S1 Fig. Confirmed and discarded cases of dengue, chikungunya and Zika in Brazil**. The plots show number of confirmed and discarded cases per epidemiological week of: dengue, from 2010 until 2017; chikungunya, from 2014 until 2017; and Zika, from 2015 until 2017.

**S1 Appendix. Summary of regression results**. VAR model for the 6-dimensional multivariate time series analysis of confirmed and discarded cases of dengue, chikungunya and Zika.

**S2 Fig. Autocorrelation function (ACF) plot of the residuals with** 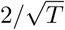 **bounds**. The plots along the diagonal are the individual ACFs for each model’s residuals, the remaining subplots show the cross-correlations between pairwise residuals. Here *T* is the sample size. The plots suggests randomness of the residuals, indicating a good fitting of the VAR model.

**S3 Fig. Probability plot and histograms for confirmed and discarded Zika model’s residuals**. The probability plot shows the unscaled quantiles of residuals versus the probabilities of a normal distribution.

**S4 Fig. Probability plot and histograms for confirmed and discarded chikungunya model’s residuals**. The probability plot shows the unscaled quantiles of residuals versus the probabilities of a normal distribution.

**S5 Fig. Probability plot and histograms for confirmed and discarded dengue model’s residuals**. The probability plot shows the unscaled quantiles of residuals versus the probabilities of a normal distribution.

## Acknowledgments

We would like to thank all the specialists for the discussions and support regarding the modeling approach.

This work is supported by the Center of Data and Knowledge Integration for Health (CIDACS) through the Zika Platform-a long-term surveillance platform for the Zika virus and microcephaly, Unified Health System (SUS) - Brazilian Ministry of Health.

## References

1. Wilder-Smith, A., Gubler, D. J., Weaver, S. C., Monath, T. P., Heymann, D. L., and Scott, T. W. Epidemic arboviral diseases: priorities for research and public health. The Lancet infectious diseases, 17(3), e101–e106. (2017).

2. Paixão ES, Teixeira MG, Rodrigues LC. Zika, chikungunya and dengue: the causes and threats of new and re-emerging arboviral diseases. BMJ global health. 2018 Jan 1;3(Suppl 1):e000530.

3. BRASIL., Ministério da Saúde. Secretaria de Vigilância em Saúde. Coordenação Geral de Desenvolvimento da Epidemiologia em Serviços. Guia de Vigilância em Epidemiológica. http://portalarquivos.saude.gov.br/images/pdf/2017/outubro/06/Volume-Unico-2017.pdf

4. World Health Organization. Global strategy for dengue prevention and control 2012-2020.

5. Teixeira Maria G., et al. East/Central/South african genotype chikungunya virus, Brazil, 2014. Emerging infectious diseases, 2015, 21.5: 906.

6. Campos GS, Bandeira AC, Sardi SI. Zika virus outbreak, bahia, brazil. Emerging infectious diseases, 2015, 21.10: 1885.

7. Ioos S, Mallet HP, Goffart IL, Gauthier V, Cardoso T, and Herida M Current Zika virus epidemiology and recent epidemics. Medecine et maladies infectieuses, 2014, 44.7: 302–307.

8. Waggoner JJ, Gresh L, Vargas MJ, Ballesteros G, Tellez Y, Soda KJ, Sahoo MK, Nuñez A, Balmaseda A, Harris E, Pinsky BA. Viremia and clinical presentation in Nicaraguan patients infected with Zika virus, chikungunya virus, and dengue virus. Clinical Infectious Diseases. 2016 Aug 30:ciw589.

9. Donalisio MR, Freitas AR, Zuben AP. Arboviruses emerging in Brazil: challenges for clinic and implications for public health. Revista de saude publica. 2017 Apr 10;51:30.

10. Diggle PJ. Time Series : a Biostatistical Introduction. Oxford: Clarendon press, 1990.

11. Lütkepohl H. New introduction to multiple time series analysis.Springer Science and Business Media; 2005 Dec 6.

12. Oliveira JF. Multivariate time series analiysis of Zika dengue and chikungunya viruses in Brazil. GitHub repository. https://github.com/Julian-sun/Multivariate-time-series-analiysis-of-Zika-dengue-and-chikungunya-viruses-in-Brazil.git

13. de Oliveira WK, de França GV, Carmo EH, Duncan BB, de Souza Kuchenbecker R, Schmidt MI. Infection-related microcephaly after the 2015 and 2016 Zika virus outbreaks in Brazil: a surveillance-based analysis. The Lancet. 2017 Aug 26;390(10097):861–70.

